# Sample Pooling as an efficient strategy for SARS-COV-2 RT-PCR screening: a multicenter study in Spain

**DOI:** 10.1101/2020.07.04.20146027

**Authors:** Adolfo de Salazar, Antonio Aguilera, Rocio Trastoy, Ana Fuentes, Juan Carlos Alados, Manuel Causse, Juan Carlos Galán, Antonio Moreno, Matilde Trigo, Mercedes Pérez, Carolina Roldán, Ma José Pena, Samuel Bernal, Esther Serrano-Conde, Gema Barbeito, Eva Torres, Cristina Riazzo, Jose Luis Cortes-Cuevas, Natalia Chueca, Amparo Coira, Juan M Sanchez-Calvo, Eduardo Marfil, Federico Becerra, María José Gude, Ángeles Pallarés, María Luisa Pérez Del Molino, Federico García

**Author notes:** These authors contributed equally to this work (Adolfo de Salazar, Antonio Aguilera). Corresponding author: Federico García.

## Abstract

**Importance:** The actual demand on SARS-CoV-2 diagnosis is a current challenge for clinical laboratories. Sample pooling may help to ameliorate workload in clinical laboratories.

**Objective:** to evaluate the efficacy of sample pooling compared to the individual analysis for the diagnosis of CoVID-19, by using different commercial platforms for nucleic acid extraction and amplification.

**Design and settings:** observational, prospective, multicentre study across 9 Spanish clinical microbiology laboratories including SARS-CoV-2 RNA testing performed in April 2020, during the first three days after acceptance to participate.

**Participants and Methods:** 3519 naso-oro-pharyngeal samples received at the participating laboratories were processed individually and in pools (351 pools) according to the existing methodology in each of the centres.

**Results:** We found that 253 pools (2519 samples) were negative, and 99 pools (990 samples) were positive; with 241 positive samples (6.85%), our pooling strategy would have saved 2167 PCR tests. For 29 pools (made out of 290 samples) we found discordant results when compared to their correspondent individual samples: in 24/29 pools (30 samples), minor discordances were found; for five pools (5 samples), we found major discordances. Sensitivity, specificity, positive and negative predictive values for pooling were 97.93%, 100%, 100% and 99.85% respectively; accuracy was 99.86% and kappa concordant coefficient was 0.988. As a result of the sample dilution effect of pooling, a loss of 2-3 Cts was observed for E, N or RdRP genes.

**Conclusion:** we show a high efficiency of pooling strategies for SARS-CoV-2 RNA testing, across different RNA extraction and amplification platforms, with excellent performance in terms of sensitivity, specificity, and positive and negative predictive values. We believe that our results may help clinical laboratories to respond to the actual demand and clinical need on SARS-CoV-2 testing, especially for the screening of low prevalence populations.

**Key points:** 

**Question:** May clinical laboratories implement sample pooling as an efficient and safe strategy for SARS-COV-2 RT-PCR screening?

**Findings:** Sensitivity, specificity, positive and negative predictive values for pooling were 97.93%, 100%, 100% and 99.85% respectively; accuracy was 99.86% and kappa concordant coefficient was 0.988.

**Meaning:** Sample pooling can be used safely at clinical laboratories, especially for the screening of low prevalence populations.

## Introduction

SARS-CoV-2 pandemic has posed an immense challenge for the National Health Systems of the affected countries, and also with regard to diagnosis. In the absence of a vaccine or effective treatment, molecular diagnosis is the only tool to contain the pandemic, identifying the transmitting infected patients, and proceed to their isolation to avoid new infections. Due to its high demand, testing opportunities may have been hampered in some scenarios, as a consequence of the lack of reagent supplies, and their limited production.

The current challenge for SARS-CoV-2 diagnosis is the great demand of testing that we are facing in the new test and trace era. Clinical laboratories must plan to increase their analytical capacity to face these new public health challenges that will allow, by means of massive analysis, to identify all infected persons, proceed with their isolation and trace their contacts. This undoubtedly constitutes a challenge due to the high number of diagnostic processes required and the limited resources available in the face of a disease with a variable incubation period, an uncertain viral dynamic (1, 2) and an unknown number of asymptomatic carriers who can transmit the infection.

Dorfman in 1943 (3) introduced the strategy of mixing samples in a single test into clinical diagnosis, and this strategy has been helpful in correctly identifying all infected individuals using fewer diagnostic tests (4,5). The diagnosis of SARS-CoV-2 infection is fundamentally based on real-time RT-PCR, which is the reference technique (6, 7). This is a robust technology with high sensitivity and specificity and has already been used in the pooling strategy of samples for the screening of HIV, HBV and HCV (8-11), where it has proven to be cost-effective and efficient in surveillance and diagnosis (detection) for prevalence below 30%, regardless of the population studied. Therefore, the combination of pooled tests and patients with low risk of infection is considered a practical and effective method to analyse large quantities of samples without compromising precision, especially when it comes to centralized models with automated systems.

In SARS-CoV-2 infection, the data on the best strategy for the detection of cases by grouping of samples and how they influence the sensitivity of the RT-PCR analysis are limited, therefore, it is necessary to investigate the effect of the number of samples, especially with commercially available assays (12-17)

Our objective in this study has been to evaluate the efficacy of sample pooling in a multicentre way compared to the individual analysis for the detection of CoVID-19 by using different commercial platforms available for genomic extraction and amplification by RT -PCR in real time.

## Materials & Methods

### Specimen collection (Individual testing)

Between March and May 2020, naso-oro-pharyngeal swabs (n = 3519) were collected from patient or health professionals and sent to the virology laboratories of the participating centres (Supplementary table 1). Samples were inactivated 1:1 in lysis buffer and processed according to the existing methodology in each laboratory (Table 1).

**Table 1.** Nucleic Acid Extraction and Amplification systems used, and number of samples studied

| Extraction | Amplification | Target | n |
| --- | --- | --- | --- |
| Maxwell® RSC Viral Total Nucleic Acid (Promega) | Viasure SARS-CoV-2 Real Time PCR (CerTest) | ORF1ab and N gene | 139 |
| m2000sp (Abbot) | Viasure SARS-CoV-2 Real Time PCR (CerTest) | ORF1ab and N gene | 410 |
| MagMAX™ Viral RNA isolation Kit (Thermo Fisher Scientific) | TaqMan 2019-nCoV Assay Kit v1 (Thermo Fisher Scientific) | N gene | 70 |
| eMAG™ (BioMeriux) | TaqMan 2019-nCoV Assay Kit v1 (Thermo Fisher Scientific) | N gene | 190 |
| STARMag™ (Seegene) | Allplex™ 2019-nCoV Assay (Seegene) | E, RdRP and N gene | 1910 |
| MagMAX™ Viral RNA isolation Kit (Thermo Fisher Scientific) | Light Mix E gene (Roche) | E gene | 240 |
| cobas® SARS-CoV-2 test (Roche) |  | E and ORF1a gene | 560 |

### Pooled analysis

Nine or ten individual samples were pooled, and screening was performed using reverse transcriptase–polymerase chain reaction targeting the same target as for individual samples.

### Performance characteristics of pooled analysis

Major discordance was defined as a negative pool result when at least one of the individual samples showed Ct values <35 for one or more SARS-COV-2 genes. Minor discordant calls were considered when at least one individual sample had a Ct value >35 in one or two of the SARS-COV-2 genes assayed and the pool scored negative.

The performance characteristics such as sensitivity, specificity, positive predictive value (PPV), negative predictive value (NPV) and relative efficiency was calculated comparing the individually analysed sample (gold standard) with respect to the sample result analysed within the pool. Statistics were performed on R Studio and Graph Pad Prism V8 software.

### Ethical approval

This study was approved by “Comité de Ética de la Investigación con medicamentos de Galicia (CEIm-G)” review board. Given the deidentified nature of testing, individual patient consent was not required for this study.

## Results

The study included 3519 samples from 11 different sites in Spain. We analyzed all samples individually and also in parallel, pooled into 353 groups of samples (342 pools of 10 samples and 11 pools of 9 samples). Two hundred and forty-one (6.85%) of the samples were positive.

We found that 253 pools, made up of 2519 samples, were negative (242 pools of 10 samples and 11 pools of 9 samples). On the other hand, we found that 99 pools, made up of 990 samples, were positive (99 pools of 10 samples). One pool, made of 10 samples, was invalid. Therefore, our pooling strategy would have saved 2167 (86%) PCR tests. A description of positive and negative pools can be found in table 2.

**Table 2.** Detailed description of the number of PCR tests performed with and without the pooling strategy

| Result | Pool | Samples Individual | Analyzed with pool strategy |
| --- | --- | --- | --- |
| Positive | 99 | 990 | 99 + 990 |
| Negative | 253 | 2519 (2420+99) | 253 |
| Invalid | 1 | 10 | 10 |
| Total | 353 | 3519 | 1352 |

Overall, 323 pools with 3219 samples showed concordant results with the individual samples analyzed (242 pools with 2229 samples negative and 99 pools with 990 samples that included at least one positive sample). For 29 pools (290 samples) we found discordant results compared to individual samples. In 24 pools minor discordances were found and in 5 pools we found major discordances. A detailed description of the discordances can be found in Table 3. The pools with minor discordances included samples typically obtained from patients who had a prior positive SARS-CoV 2 test and were submitted for PCR testing at least 20 days after to evaluate RNA clearance

**Table 3.**
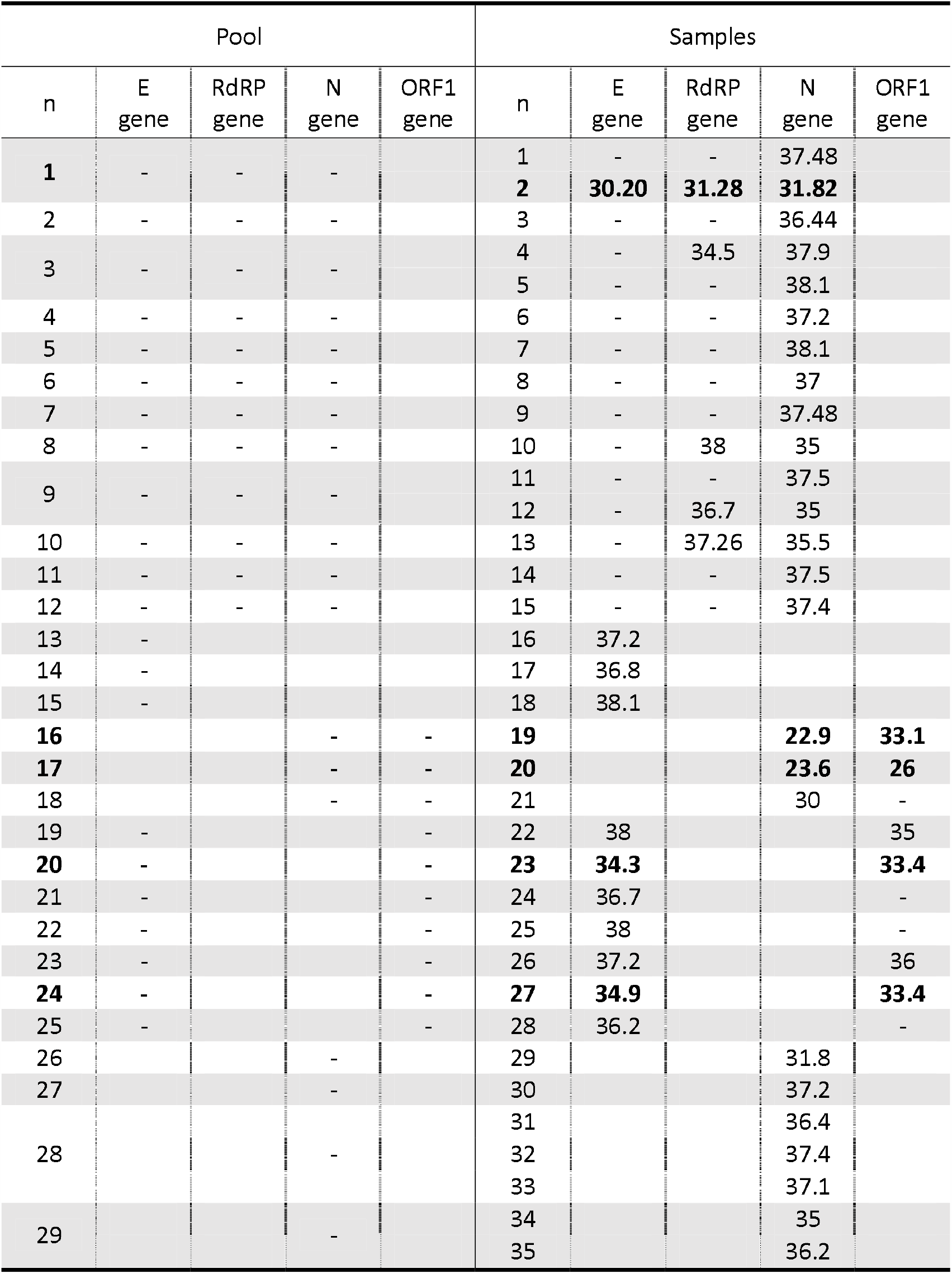
Discordant results between pools and individual samples. Major discordances are highlighted in bold.

When only major discordances were considered for the analysis, sensitivity, specificity, positive and negative predictive values for pooling were 97.93%, 100%, 100% and 99.85% respectively. Accuracy was also 99.86% and kappa concordant coefficient was 0.988. These data are detailed in Table 4.

**Table 4.** Performance characteristics of Pooled analysis.

| Major discordance |  | Individual PCR (Reference) |  |
| --- | --- | --- | --- |
|  |  | Positive | Negative |
| Pool PCR | Positive | 236 | 0 |
|  | Negative | 5 | 3268 |

| Statistic | Value | 95% CI |
| --- | --- | --- |
| Sensitivity | 97.93% | 95.23% to 99.32% |
| Specificity | 100.00% | 99.89% to 100.00% |
| Positive Predictive Value | 100.00% |  |
| Negative Predictive Value | 99.85% | 99.64% to 99.94% |
| Accuracy | 99.86% | 99.67% to 99.95% |
| kappa (k) | 0.988 |  |

When all discordances were considered for the analysis, sensitivity, specificity, positive and negative predictive values for pooling were 85.48%, 100%, 100% and 98.94% respectively. Accuracy was 99% and kappa concordant coefficient was 0.916. These data are detailed in Table 5.

**Table 5.** Performance characteristics of Pooled analysis.

| All discordance |  | Individual PCR (Reference) |  |
| --- | --- | --- | --- |
|  |  | Positive | Negative |
| Pool PCR | Positive | 206 | 0 |
|  | Negative | 35 | 3268 |

| Statistic | Value | 95% CI |
| --- | --- | --- |
| Sensitivity | 85.48% | 80.39% to 89.67% |
| Specificity | 100.00% | 99.89% to 100.00% |
| Positive Predictive Value | 100.00% |  |
| Negative Predictive Value | 98.94% | 98.57% to 99.22% |
| Accuracy | 99.00% | 98.62% to 99.30% |
| kappa (k) | 0.916 |  |

An in-depth analysis of the Ct results was performed to check the pooling effect on Ct differences. Of the 99 positive pools, 42 were positive pools of 10 samples with 1 positive sample each. These samples and pools were tested for the E, RdRP and N genes (Allplex ™ 2019-nCoV Assay and analysed by the 2019-nCov Seegene Software). The difference in Ct results between pool and positive sample can be seen in figure 1. Interestingly, we found that the most sensitive target after pooling was the N gene (41 pools positive), followed by the RdRP gene (33 pools positive); the E gene (27 pools positive) showed the least sensitivity after pooling samples.

**Figure 1.**
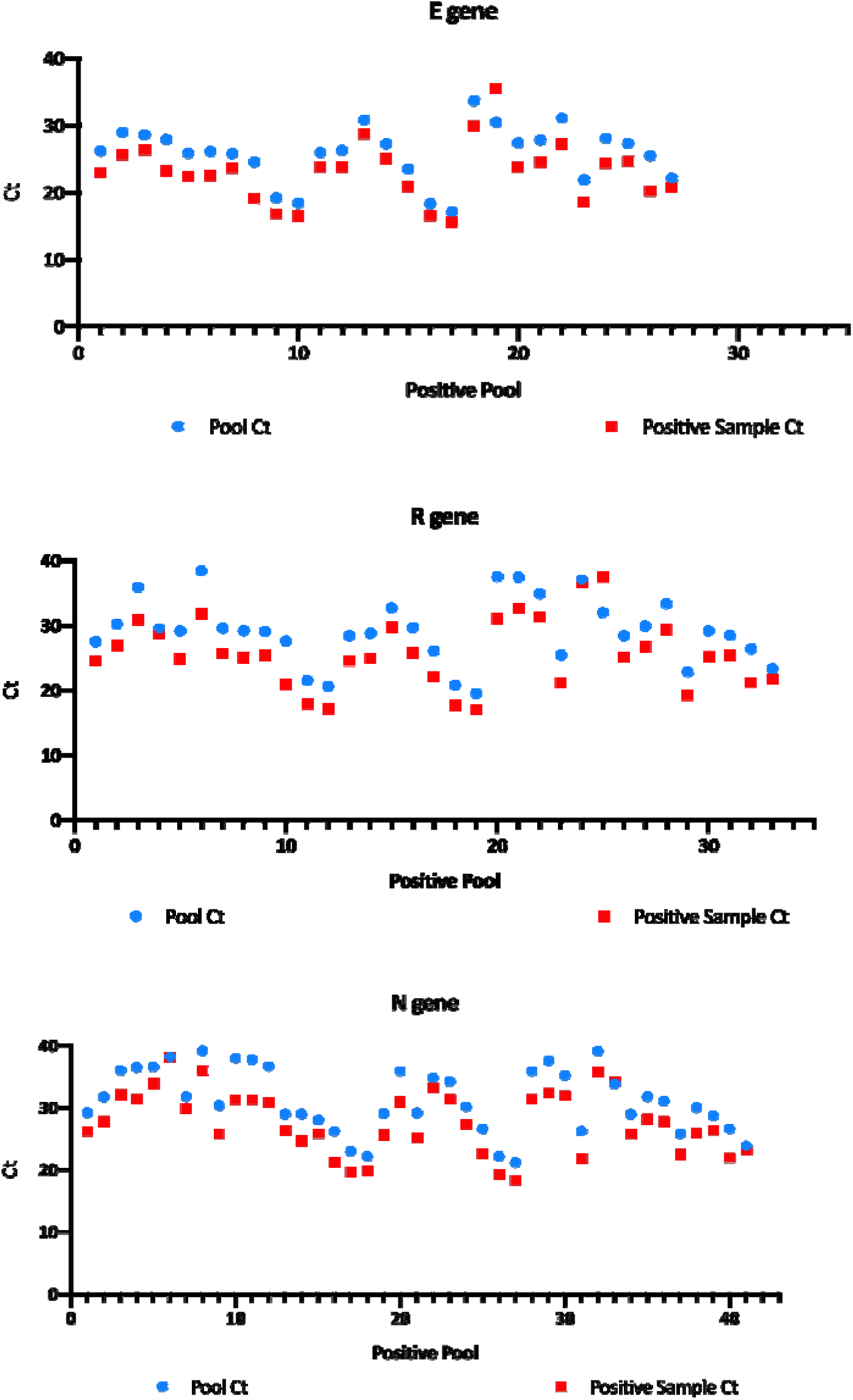
Pooling effect on Ct results of the specific E, RdRP and N genes from the Allplex ™ 2019-nCoV Assay (Seegene)

## Discussion

The increasing demand for testing of SARS-CoV-2 by RT-PCR requests new strategies that can accommodate the great number of tests that clinical laboratories need to process. Nucleic acid testing allows for the use of sample pooling strategies, that have been successfully used for a broad number of pathogens. Here we report on the high efficiency of pooling strategies for SARS-CoV-2 RNA testing, across different RNA extraction and amplification platforms, with excellent performance in terms of sensitivity, specificity, and positive and negative predictive values. We believe that our results may help clinical laboratories to respond to the actual demand and clinical need on SARS-CoV-2 testing and may serve as an experience for future needs in clinical microbiology.

Spain with more than 230,000 confirmed cases of CoVID-19 and more than 27,000 deaths at the end of May 2020 is one of the most affected countries in the world by the pandemic caused by the new coronavirus 2019 (SARS-CoV-2) according to data from the Centers for Systems Science and Engineering (CSSE) at Johns Hopkins University (JHU), and the ECDC (https://github.com/EU-ECDC/PooledPrevalence). Pooling strategies may allow to increase the analytical capacity of clinical laboratories and hence will allow to face this new public health challenge. Although sample pooling strategy works for other pathogens that are diagnosed by RT-PCR, given the novelty of CoVID-19 and its impact in public health, it seems wise to be cautious and to validate this strategy before being able to recommend it. For SARS-CoV-2 infection, there is still limited data in the literature regarding surveillance and detection strategies by grouping samples in the quality measures of RT-PCR analysis (12-17)

In our study we have evaluated the efficacy of sample pooling in a multicentre way compared to the individual analysis for the detection of CoVID-19 by using different commercial platforms available for genomic extraction and amplification by RT-PCR in real time. Within a 6.8% positive rate, we have obtained excellent numbers in sensitivity, specificity, and positive and negative predictive values, both in a scenario for which only major discordances were considered (97.98%, 100%, 100%, 99.85%, respectively) and also when minor discordances were counted (85.48%. 100%, 100% and 98.94%).

As expected, and as other authors have also shown (18), the dilution of samples in our pooling strategy resulted into a median loss of 2.87 CTs for E gene, 3.36 CTs for RdRP gene and 2.99 CTs for N gene. This drop in the sensitivity was responsible for most of the discordances found in our study, that were mainly observed for samples with the lowest positivity signals, always with CTs very close to 40, and very frequently in only one gene of the two-three that were included in the tests. Although special attention on RT-PCR false negative results must be paid (19), it is also known that most of the positive results obtained from just one gene targeted and with CTs >35 correspond to non-viable/noninfectious particles that are still detected by RT-PCR (20).

Our study’s main limitation was the variability upon extraction and amplification methods used, and the number of samples included in the different pools tested; however, this limitation in the design may turn into its main strength given the consideration that even in this scenario our results were excellent. In the sample pooling strategy, it is a priority to determine the group size in which the maximum precision is maintained in the analysis performed, since, due to the dilution of the sample, this procedure can decrease the sensitivity of the molecular assays of RT- Real-time PCR “Optimization of group size in pool testing strategy for SARS-CoV-2: A simple mathematical model” (21). For this reason, before systematically implementing a sample grouping strategy, it is important to consider these characteristics (detection limit, sensitivity and specificity of the test) together with the expected prevalence; in this regard, there are already applications that allow it to be calculated (https://www.chrisbilder.com/shiny/). In addition, the main advantages of the pooling strategy are that it allows using the same standard protocols of commercial reagents, with no need of additional training, equipment or materials, and consequently it can be implemented immediately to expand the detection and surveillance capabilities of CoVID-19.

It should be noted that pooling as a screening strategy will not completely eliminate the need for individual diagnostic tests, which will be essential when community transmission intensifies. In our study, even in the setting of a 6.86% prevalence, out of 3617 samples analysed, we would have saved a total of 2456 PCR tests, with a great saving in time, costs and personnel. Within the current epidemiological situation, in which prevalence has significantly decreased but the high demand on PCR testing continues, this efficiency measures would help clinical laboratories in alleviating the workload in order to prepare for future outbreaks

In summary, we show a high efficiency of pooling strategies for SARS-CoV-2 RNA testing, across different RNA extraction and amplification platforms, with excellent performance in terms of sensitivity, specificity, and positive and negative predictive values. As we believe our findings may be essential to expand clinical laboratories capabilities in the very near future, we recommend to validate this strategy in each specific setting of extraction and amplification reagents before its introduction for clinical management, specially to ensure that sensitivity of the assay and especially the false negative rates are acceptable.

## Data Availability

All data relevant to the study are included in the article or uploaded as supplementary information

## Conflict of interest

The authors declare that they have no conflicts of interest.

## Tables

**Supplementary table 1.** Number of samples analyzed in each laboratory

| Hospital | n |
| --- | --- |
| Hospital Universitario Clínico San Cecilio | 970 |
| Complejo Hospitalario Universitario de Santiago | 860 |
| Hospital Universitario de Jerez | 410 |
| Hospital Universitario Reina Sofia | 400 |
| Hospital Universitario Ramon y Cajal | 350 |
| Hospital Universitario Lucus Augusti | 200 |
| Complejo Hospitalario Universitario de Pontevedra | 139 |
| Hospital Universitario Virgen de las Nieves | 110 |
| Hospital Universitario de Jaen | 80 |
| Total | 3519 |

## Notes

### Competing Interest Statement

The authors have declared no competing interest.

### Funding Statement

No external funding was received for this study.

### Author Declarations

This study was approved by Comite de Etica de la Investigacion con medicamentos de Galicia (CEIm-G) review board. Given the deidentified nature of testing, individual patient consent was not required for this study.

